# Pentoxifylline and Covid-19: A Systematic Review

**DOI:** 10.1101/2020.09.14.20194381

**Authors:** Diego Ramonfaur, Carlos A. González-Assad, José G. Paredes-Vázquez

## Abstract

At more than 10 months after the first case of COVID-19 was documented, the understanding of the pathogenesis of this viral illness is growing on a daily basis. A massive pro-inflammatory response on infected individuals involving several cytokines seems to play a key role on disease. As a result, therapeutic efforts have focused on anti-inflammatory strategies to ameliorate the disease, in sight of a lack of a truly effective anti-viral agent. Pentoxifylline (PTX) has been proposed by multiple authors as a potential therapeutic ally, targeting a variety of mechanisms as it has been shown to have antiviral, anti-inflammatory and hemodynamic effects. Importantly, anti-inflammatory effects center on down-regulation of cytokines such as interleukins and tumor necrosis factor. In pre-pandemic studies, PTX has demonstrated to change the clinical course of inflammatory diseases such as acute respiratory distress syndrome, which is a hallmark of severe COVID-19. Researchers agree it is pertinent to experimentally evaluate the effect this drug has on COVID-19 patients. The objective of this review is to summarize all the proposed mechanisms by which PTX may aid in the treatment of COVID-19, as well as prevent its deadly complications. Our interpretation of the literature is that the benefits PTX may bring to a patient with COVID-19 outweigh the risks this drug might pose on them. As a result, there is consensus regarding the evaluation of PTX in further experimental studies to better characterize its effects on COVID-19 patients.

## INTRODUCTION

In late 2019, the Severe acute respiratory syndrome coronavirus 2 (SARS-CoV-2) emerged in Wuhan, China, presumably as a zoonotic infection originating from bats.(1) This virus, which causes the coronavirus disease 19 (COVID-19), rapidly expanded worldwide, as travel restrictions failed to optimally control the spread. As of January 21^st^, the first case of COVID-19 was identified in the United States. Recent experience with viral respiratory epidemics with similar characteristics was limited to SARS and MERS outbreaks, as well as the 2009 H1N1 pandemic. However, it has a slightly higher rate of transmission than these viruses, and shares almost 80% of its genome with SARS-CoV.(1,2) As major pharmaceutical companies and research efforts race to find an effective therapeutic agent, many anti-viral medications were quickly tested and discarded from the list of potential medications with the exception of remdesivir, which has demonstrated a reduction in symptom duration, but is still reserved for severe cases. (3). It has been well described that a subset of patients may develop coagulopathy, as a result of the cytokine storm mediated amid an aggressive inflammatory response to the SARS-CoV-2, ultimately leading to complications like acute respiratory distress syndrome (ARDS).(4) As a result, investigators have proposed alternative drugs with multiple mechanisms of action in light of finding a lower rate of complications from this disease, and potentially lower the mortality. Pentoxifylline (PTX) is a methylxanthine derivative, currently used for peripheral vascular disease, with anti-inflammatory and immunomodulatory properties(5–7). Moreover, the main pharmacodynamic properties of this drug are aimed at improving circulation by increasing erythrocyte deformability.(8) Additionally, a safe side effect profile, and a low cost make PTX a strong candidate to be considered as an aid in the treatment for high risk COVID-19 patients. This drug was initially proposed as an alternative treatment for the SARS outbreak in 2003. Subsequently, there has been some attention to this drug as the COVID-19 pandemic began to spread. Accordingly, no published study has analyzed the clinical impact of utilizing this drug as a potential therapeutic agent. The purpose of this study is to systematically gather all the latest evidence on pentoxifylline as a potential treatment for COVID-19 and analyze the feasibility of further trials as well as trends in treatment strategies.

## METHODS

Currently, no systematic review of the literature looking at the use of pentoxifylline as a potential treatment for COVID-19 has been published. The PROSPERO database was revised, and no study of this characteristics was found. We sought to identify all scientific literature analyzing the potential benefits of pentoxifylline against COVID-19. We queried Pubmed and Google scholar search engines utilizing the following commands: (Pentoxifylline AND COVID), (Pentoxifylline AND Coronavirus), and (Pentoxifylline AND SARS). For obvious reasons, only publications from 2020 were considered. There was no language limitation. Only published manuscripts were eligible. Information regarding potential mechanisms of action of Pentoxifylline against COVID-19 and its complications was extracted. This systematic review is structured using the PRISMA guidelines.

## RESULTS

A total of 11 studies were found using the described methodology. One study which was a clinical trial proposal preprint was excluded. We identified a total of 8 research items eligible for our review which are summarized in table 1. No experimental study was found.

**Table 1:**
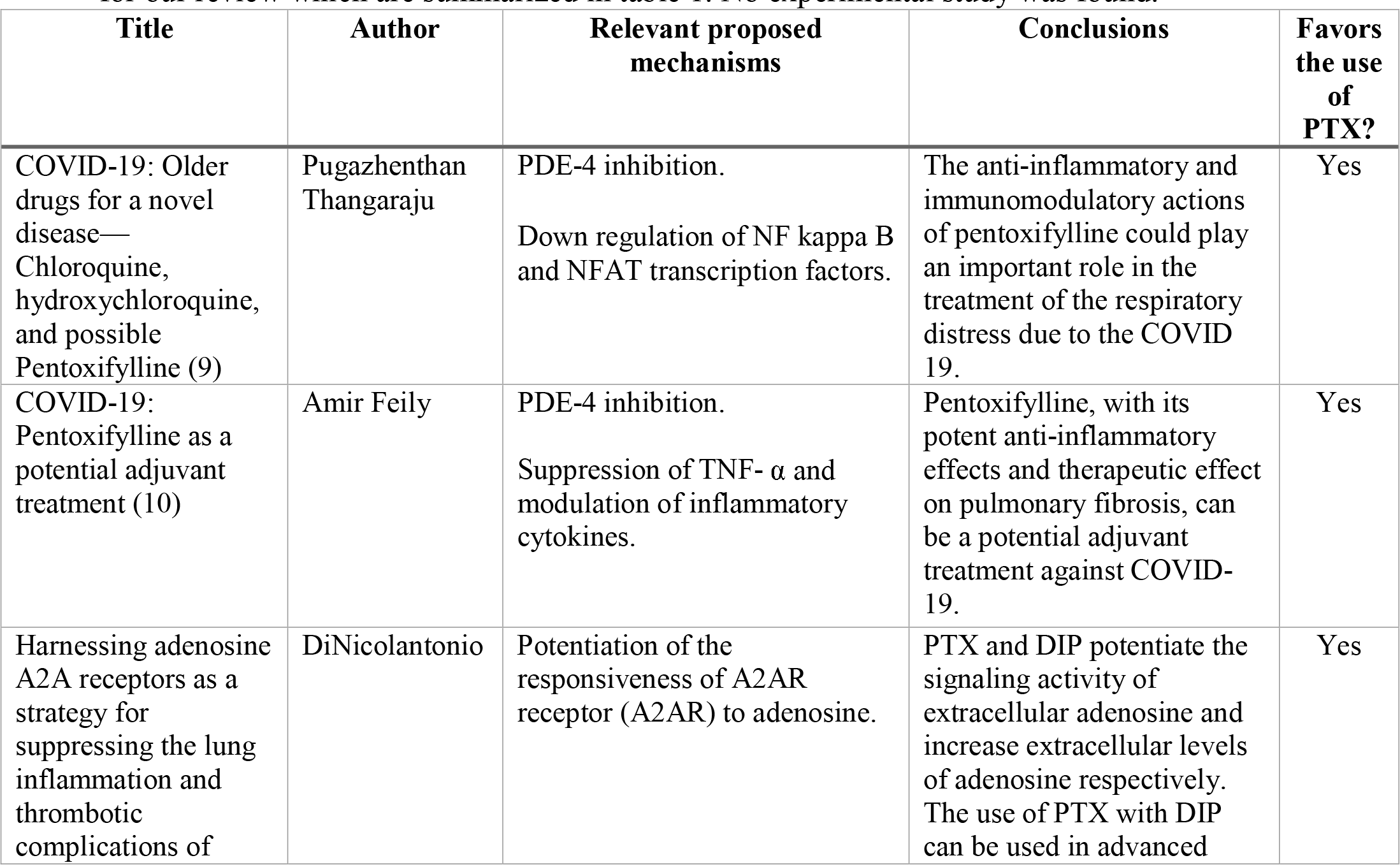
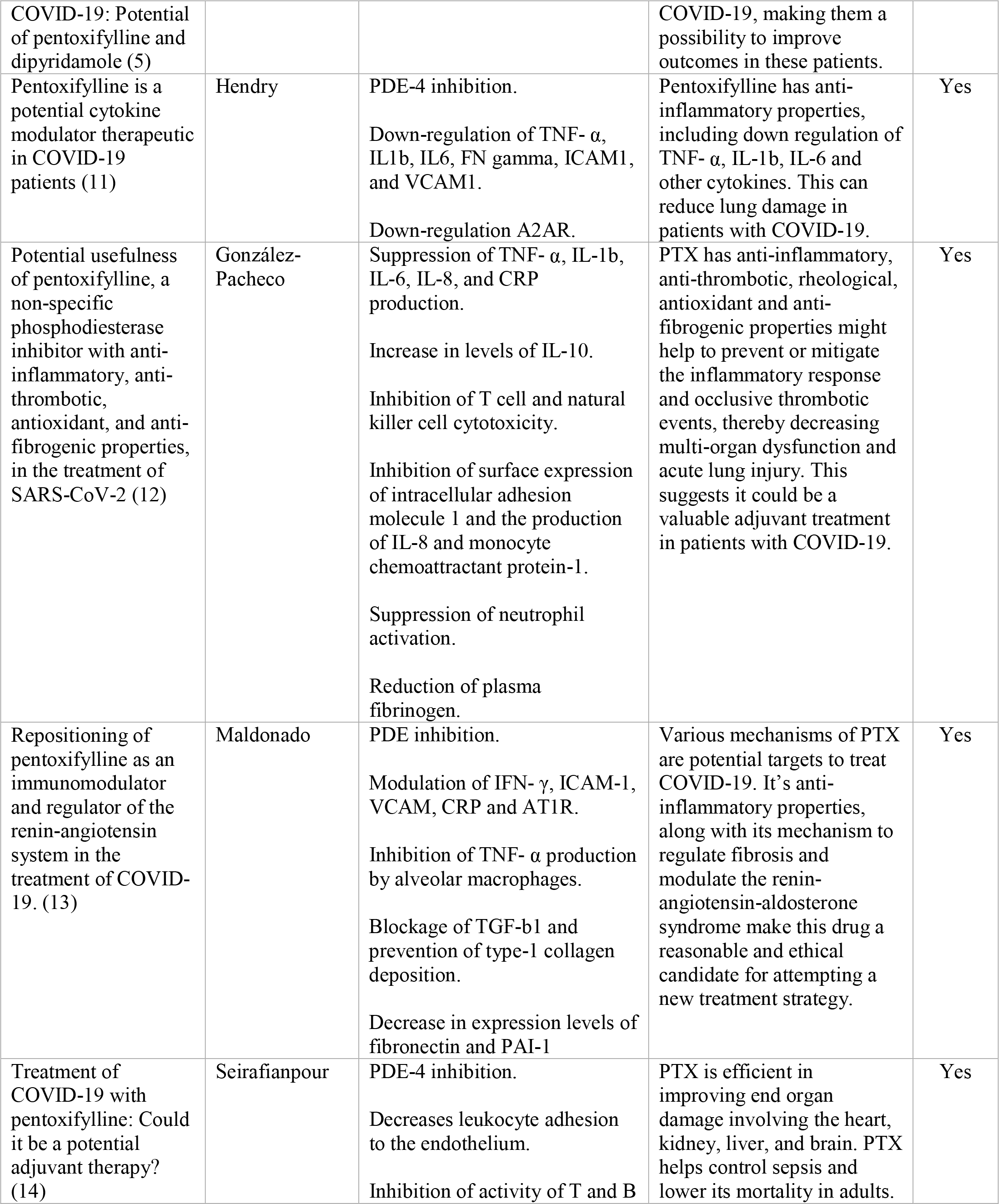
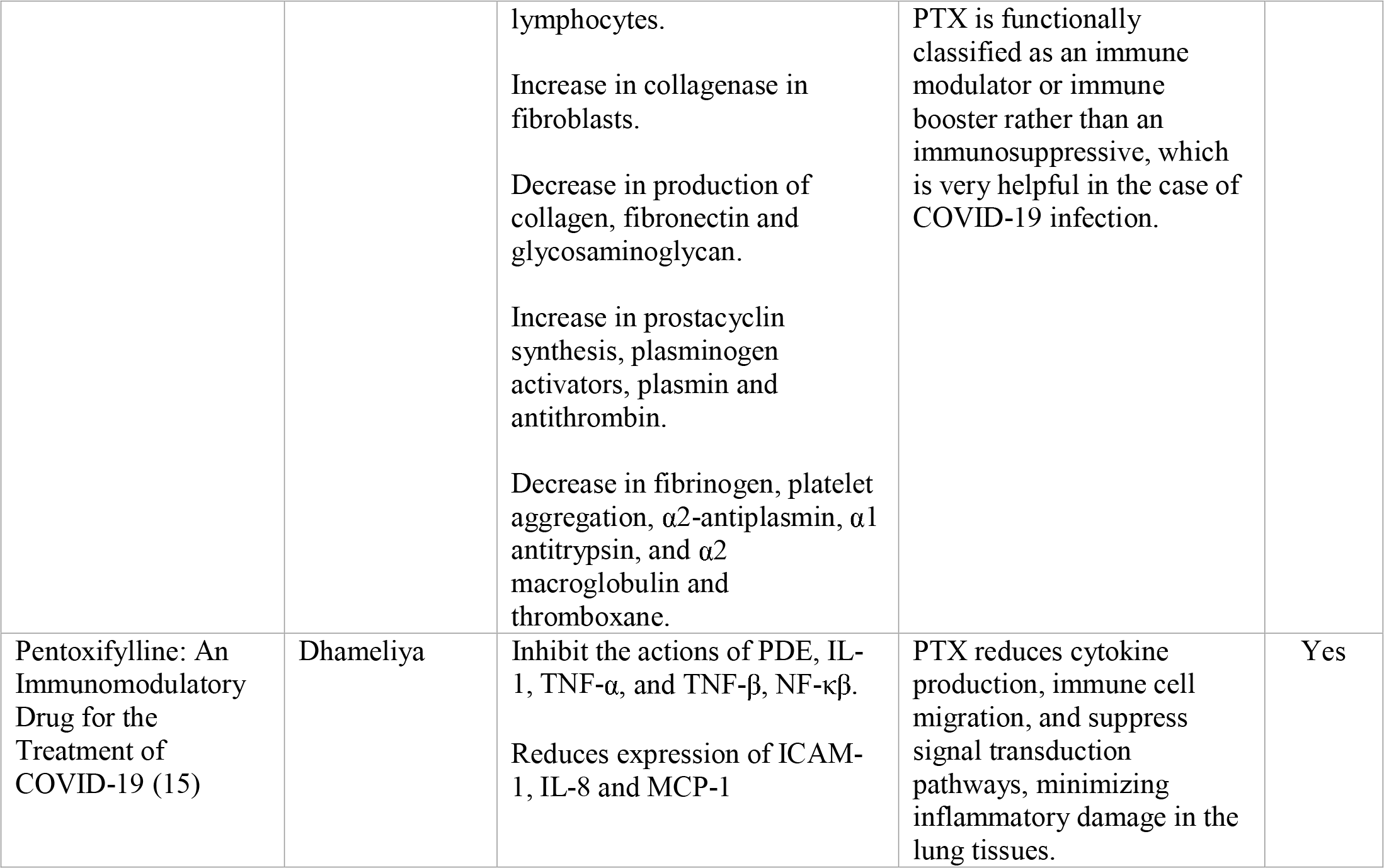
Summary of articles proposing pentoxifylline as a potential treatment strategy for COVID-19 patients. PDE-4: Phosphodiesterase-4, TNF-α:Tumor Necrosis Factor alpha, TNF-β:Tumor Necrosis Factor beta, DIP: Dipyridamole, IL-1: Interleukin-1, IL-1b: Interleukin-1 beta, IL-6: Interleukin-6, IL-8: Interleukin-8, IL-10: Interleukin-10, NF-κβ Nuclear Factor kappa beta, ICAM-1: Intercellular adhesion molecule 1, VCAM-1: Vascular cell adhesion molecule 1, NFAT: Nuclear factor of activated T-cells, MCP-1: Monocyte Chemoattractant Protein-1, PAI-1: Plasminogen activator inhibitor-1, TGF-β1: Transforming growth factor beta 1, IFN-y: Interferon gamma, CRP: C-reactive protein, AT1R: Angiotensin II type 1 receptor, A2AR: Adenosine A2A receptor, STAT3: Signal transducer and activator of transcription 3

## DISCUSSION

COVID-19 illness is a virus-mediated syndrome which may develop a host cytokine storm. This pro-inflammatory response plays a key role in the damage viral infections cause on a host and may correlate with worse outcomes. There is growing evidence suggesting a subset of these patients are at risk of developing devastating complications such as myocarditis, acute respiratory distress syndrome (ARDS), acute kidney injury, and coagulopathy, among others. These complications are associated with a significant rate of morbidity and mortality, especially in patients with comorbidities, non-white race or in poverty. (16–19) Attempts to find an effective antiviral drug have been futile. The conventional antiviral drug, remdesivir seems to have a potential benefit, although there is mixed evidence regarding its effectiveness in reducing mortality and length of stay. (3,20) It has been shown that most of the dangerous complications are due to an overt inflammatory response by the host, rather than viral cytotoxicity by itself. A certain degree of inflammation is necessary to eliminate any kind of infection. However, it has been shown that persistent elevations of pro-inflammatory cytokines are associated with worst outcomes in patients with ARDS. (21) SARS-CoV-2 induces excessive and prolonged cytokine responses in some individuals, known as the cytokine storm. It is this exaggerated response, that may trigger multiple organ dysfunction. (4) As a result, immunomodulatory and anti-inflammatory drugs have received attention as potential mitigators of severe COVID-19. Some specific drugs immune modulator drugs with diverse mechanisms of action have the ability to attenuate the cytokine storm. Particularly, steroids such as dexamethasone, and interleukin antagonists like tocilizumab, and anakinra, among others, have received attention as treatment strategies for the hyperinflammatory response. (22,23) A recent study by the RECOVERY collaborative group, found a mortality reduction of 17% in the treatment arm with daily 6mg of dexamethasone for 10 days in hospitalized COVID-19 patients. (24)

Pentoxifylline has demonstrated anti-inflammatory, anti-thrombotic, immunomodulatory, hemodynamic, and antiviral properties which makes it a valuable candidate for consideration as alternative or complementary treatment for patients with moderate to severe symptoms. Modulation of TNF-α, IL1, IL6, ICAM1, VCAM1 and TNF, as well as inactivation of PDE and prevention of platelet aggregation are among the proposed mechanisms that could aid as treatment options for moderate to severe COVID-19 patients.(5,6,8,13,25,26) PTX has proven to be useful in patients with ARDS, from different etiologies and to improve pulmonary function and control pulmonary fibrosis. (27–29), Furthermore, the potential of pentoxifylline as a therapeutic agent is warranted as there is little to no downside in using this drug because of a safe adverse effect profile and a low cost.

Amid an urge for an effective treatment with a positive effect on morbidity and mortality, hundreds of drugs have been proposed as treatment strategies. This review summarizes the best available evidence showing potential benefits of using PTX as a broad-spectrum treatment mainly aimed at preventing and treating the pro-inflammatory complications of COVID-19. Limitations in our study include that no statistical analysis could be made, as no tangible evidence has been published to date.

## CONCLUSION

Our interpretation of the available literature is that there is ample evidence suggesting a wide portfolio of mechanisms through which PTX may be beneficial in the treatment of COVID-19. As a result, we believe PTX has a well-suited profile to prevent and treat potential COVID-19-associated complications. Moreover, it’s low cost and minimal adverse effects make it a safe drug to consider for pilot clinical trial studies. We encourage researchers to study the clinical benefit of this drug in moderate to severe COVID-19 patients.

## Data Availability

N/a

